# Baseline tumor-infiltrating lymphocyte patterns and response to immune checkpoint inhibition in metastatic cutaneous melanoma

**DOI:** 10.1101/2023.11.27.23299053

**Authors:** Isabella A.J. van Duin, Mark Schuiveling, Laurens S. ter Maat, Wouter A.C. van Amsterdam, Franchette van den Berkmortel, Marye Boers-Sonderen, Jan Willem B. de Groot, Geke A.P. Hospers, Ellen Kapiteijn, Mariette Labots, Djura Piersma, Anne M.R. Schrader, Gerard Vreugdenhil, Hans Westgeest, Mitko Veta, Willeke A.M. Blokx, Paul J. van Diest, Karijn P.M. Suijkerbuijk

## Abstract

**Introduction:** The presence of tumor-infiltrating lymphocytes (TILs) in melanoma has been linked to survival. Their predictive capability for immune checkpoint inhibition (ICI) response remains uncertain. Therefore, we investigated the association between treatment response and TILs in the largest cohort to date and analyzed if this association was independent of known clinical predictors of response.

**Methods:** In this multicenter cohort study, patients who received first-line anti-PD1 ± anti-CTLA4 for advanced cutaneous melanoma were identified. TILs were scored as absent, non-brisk or brisk on hematoxylin and eosin (H&E) slides of primary melanoma and pre-treatment metastases. Scoring systems evaluating the infiltration and intensity patterns (‘MIA-score’) and the percentage of stromal TILs were also evaluated. The primary outcome was objective response rate (ORR), with PFS and OS being secondary outcomes. Univariable and multivariable logistic regression and Cox proportional hazard regression analyses were performed, adjusting.for age, sex, disease stage, ICI type, BRAF mutation, lactate dehydrogenase (LDH) level and WHO performance score.

**Results:** Metastatic melanoma specimens were available for 650 patients and primary specimens from 565 patients.. No association was found between TILs in primary melanoma specimens and response. In metastatic specimens, patients with non-brisk TILs (aOR 1.56, 95% CI 1.06-2.29) and brisk TILs (aOR 3.28, 95% CI 1.72-6.56) had a higher probability of response, longer median PFS (9.2 and 19.4 vs. 6.5months [p=0.009]) and median OS (49.5 and 40.9 vs 21.3 months [p=0.007]) when compared to absent TILs. Similar results were noted using the MIA- and stromal TIL scores.

**Conclusion:** In advanced melanoma patients, TIL patterns on H&E slides of pre-treatment metastases are associated with ICI response. This is independent of known clinical predictors. TILs are easily scored on readily available H&Es, which facilitates the use of this biomarker for ICI outcomes in clinical practice.

## Introduction

Immune checkpoint inhibition (ICI) has significantly improved the prognosis for patients with advanced cutaneous melanoma. Five-year overall survival (OS) rates exceed 40% for anti-PD-1 monotherapy and 50% for combination therapy with anti-CTLA-4.^1^ However, half of patients fail to respond to ICI treatment.^2^ Unfortunately, they can still experience the potentially severe side effects.27/11/2023 14:15:00 Also, ICI treatment costs are high, imposing a substantial burden to the health care system.^4^ Predicting who will respond to treatment and who will not is currently hindered by the lack of biomarkers with sufficient predictive value.

Several studies have identified tumor-infiltrating lymphocytes (TILs) as a favorable prognostic and predictive factor in melanoma, even before the era of ICI.^5,6^ Given the recent clinical successes of immune system activation in treating advanced melanoma, there is renewed interest in the presence of TILs as a potential biomarker.^7^ TILs can refer to lymphocytes present both within tumor nests and within the surrounding stroma.

In melanoma histopathology, three scoring systems have been used to score TILs on haematoxylin & eosin (H&E)-stained slides. The most widely validated and used scoring system is the one proposed by Clark *et al* in 1989, which classifies TILs on an increasing scale as absent, non-brisk or brisk.^8^ Later, Azimi *et al* proposed a more refined four-tier TIL scoring system in primary melanoma (“MIA-score”), combining pattern of infiltration (focal, multifocal, diffuse) with intensity (absent, mild, moderate, or marked) attempting to score TILs more precisely.^9^ Recently, a quantitative scoring of TILs was proposed by Hendry *et al* based on counting stromal TILs within the borders of the invasive tumor.^10^ This method was applied in both the primary and metastatic setting. Although this “stromal score” method has not been validated yet, stromal TILs might hold prognostic and/or predictive value.

The majority of studies that were conducted in ICI-treated patient cohorts have focused on scoring TILs using immunohistochemistry, flow cytometry or genomics.^11,12^ Although promising, this limits the implementation of TIL involvement as a biomarker in routine practice, since these techniques could be costly and complex. Studies that investigated the association between TIL score on H&E slides and outcomes in advanced melanoma have typically involved small subgroups of <150 ICI-treated patients.^13,14^ Additionally, the majority of these studies have focused on scoring TILs in primary melanomas, further limiting the potential of TIL involvement as a biomarker in advanced melanoma. H&E-staining forms an integral part of routine pathology procedures and, consequently, represents an existing and widely available resource for nearly all patients. Therefore, assessing TILs on H&E-stained slides could yield important biomarker information which, if feasible, can be implemented easily in day-to-day practice.

In this largest study to date, we evaluated the three TILs scoring methods on H&E-stained slides obtained from both primary and pre-treatment metastatic specimens of patients with advanced melanoma undergoing ICI-treatment. The objective was to investigate the predictive value of these TIL scores for response to ICI therapy.

## Methods

### Patients

Patients were retrospectively identified from high quality registry data from ten participating centres in the Netherlands.^15^ Patients were included if above 18 years of age and treated with first-line anti-PD1 monotherapy or anti-PD1 and anti-CTLA4 for irresectable stage IIIC or stage IV after January 1, 2016. The patient’s stage of disease was based on the 8^th^ edition of the AJCC melanoma staging system.^16^ Patients with less than 6 months of follow-up were excluded from the analysis.

### Slide selection and TIL assessment

For each patient, one representative H&E-stained slide was selected from both the primary melanoma and metastasis. Some patients had only primary melanoma or metastatic specimens available. In cases with multiple primary melanomas, the selection process prioritized the melanoma with the highest Breslow thickness and the most suspicious location in terms of regional lymph nodes involvement. Among patients with multiple specimens from metastatic sites, the specimen closest to the date of treatment initiation was selected. All selected slides were scanned with a Nanozoomer XR C12000-21/-22 (Hamamatsu Photonics, Hamamatsu, Shizuoka, Japan) at 40× magnification with a resolution of 0.22 µm per pixel. TIL scoring was performed by authors IAJD and MS, under the supervision of two experienced pathologists (WAMB and PJvD), after training and a consensus meeting. TILs were scored according to the scoring system proposed by Clark *et al* (‘Clark score’)^8^, the scoring system proposed by the Melanoma Institute of Australia (‘MIA score’)^9^ and the semi-quantitively approach which estimates the percentage of TILs in the tumoral stroma as proposed by Hendry *et al* (‘stromal score’)^10^. The stromal score was also assessed as a categorical variable. The cutoff points for these groups were based on tertiles of the metastatic stromal score, so that each group represented a roughly equal number of patients. For a small proportion of specimens, it was not possible to score the stromal score, because of the absence of tumoral stroma (for example, if the slide contained only a few tumor cells). Average scoring time was less than five minutes depending on the interprebility of the specimen. For a more detailed description of the TIL scoring systems, see supplementary tables 1-3.

### Patient outcomes

Response evaluation was determined by the treating physician and was based on the Response Evaluation Criteria in Solid Tumors, version 1.1.^17^ Responses were defined as complete response (CR), partial response (PR), stable disease (SD) or progressive disease (PD, including melanoma-related death before first response assessment. The primary outcome was objective response, defined as the best overall response of partial or complete response. Secondary outcomes were progression-free survival (PFS) and overall survival (OS).

### Statistical analysis

We used descriptive statistics to describe the study population. This included medians and interquartile intervals (IQI) for continuous variables, and percentages and frequencies for categorical variables. Cohen’s kappa coefficients were used to determine interobserver agreement for the categorical TIL scores, intra-class correlation was used for the stromal score. For the change in TIL score between paired primary and metastatic specimens, McNemar tests and intra-class correlation were used. For the relation with categorical variables and response, Chi-square tests were used. The associations between continuous variables and response were assessed using Mann-Whitney U tests. Univariable and multivariable analyses were performed with logistic regression and Cox proportional hazards regression. The proportional hazards assumption was evaluated using Schoenfeld residuals and the assumptions were met for each TIL score. In the multivariable analysis complete case analysis was performed. For follow-up data and patient outcomes, we performed survival analysis using the (reversed) Kaplan-Meier method and logrank test to assess differences in PFS and OS between groups. Analyses were performed using R statistical software (Version 4.2.2 with package survival version 3.5.0).

## Results

### Patient characteristics and outcomes

Of the 1346 eligible patients, 49 patients were excluded because of missing outcome data. Of the remaining 1297 patients, primary melanoma specimens were available for 565 patients and metastatic specimens were available for 650 patients (Figure 1). Patient characteristics of included patients are shown in table 1 and compared to those of excluded patients in supplementary table 4. In both cohorts, most patients were male, had normal LDH levels, and were above 65 years of age. In patients with a primary specimen available, TILs were more often present compared to those with a pre-treatment metastatic specimen available. Lymph nodes were identified as the most common site of origin for the majority of the metastatic specimens (supplementary table 5). The median follow-up duration in our cohort was 28 months with a median PFS of 8.4 months and a median OS of 29.1 months. The objective response rate (ORR) to ICI was similar in patients with primary melanoma specimen available and patients with metastatic specimen available (53% and 54%, respectively).

**Figure 1.**
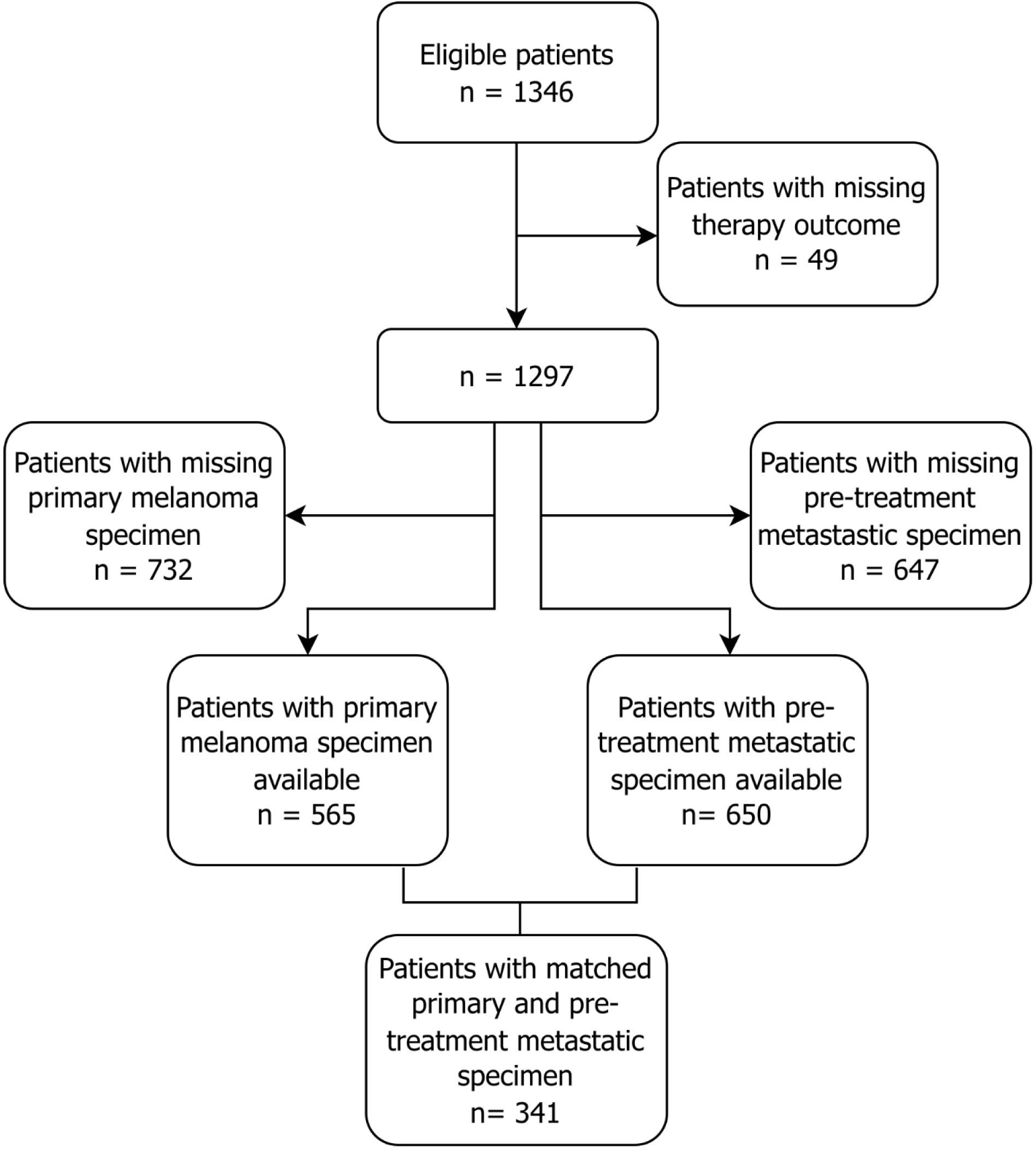
Flowchart of the study population.

**Table 1.** Patient characteristics of 565 ICI-treated patients with primary melanoma specimen available, and 650 ICI-treated patients with pre-treatment metastatic specimen available. Abbreviations: IQI, interquartile interval; LDH, Lactate dehydrogenase; TIL, tumor-infiltrating lymphocytes; ULN, upper limit of normal; WHO, World Health Organization.

|  | Primary melanoma specimen available (N=565) | Metastasis specimen available (N=650) |
| --- | --- | --- |
| **Age (years)** |  |  |
| Median [IQI] | 68 [57-75] | 66 [57-74] |
| **Sex** |  |  |
| Female | 195 (34.5%) | 228 (35.1%) |
| Male | 370 (65.5%) | 422 (64.9%) |
| **WHO Performance status** |  |  |
| WHO 0 | 262 (48.2%) | 285 (45.8%) |
| WHO 1 | 236 (43.4%) | 291 (46.8%) |
| WHO 2-4 | 46 (8.5%) | 46 (7.4%) |
| Missing | 21 | 28 |
| **Stage of disease** |  |  |
| Unresectable IIIC | 40 (7.4%) | 50 (8.1%) |
| M1a | 37 (6.8%) | 44 (7.1%) |
| M1b | 78 (14.4%) | 81 (13.1%) |
| M1c | 262 (48.3%) | 276 (44.5%) |
| M1d | 125 (23.1%) | 169 (27.3%) |
| Missing | 23 | 30 |
| **BRAF V600 Mutation** |  |  |
| Wildtype | 372 (74.1%) | 404 (70.3%) |
| Mutant | 130 (25.9%) | 171 (29.7%) |
| Missing | 63 | 75 |
| **LDH levels** |  |  |
| Not elevated | 362 (64.6%) | 413 (64.4%) |
| 1-2x ULN | 149 (26.6%) | 180 (28.1%) |
| >2x ULN | 49 (8.8%) | 48 (7.5%) |
| Missing | 5 | 9 |
| **Type of systemic therapy** |  |  |
| Anti-PD1 | 363 (64.2%) | 401 (61.7%) |
| Ipilimumab & Nivolumab | 202 (35.8%) | 249 (38.3%) |
| **Clark TILs score** |  |  |
| Absent | 138 (24.4%) | 337 (51.8%) |
| Non-brisk | 343 (60.7%) | 247 (38.0%) |
| Brisk | 84 (14.9%) | 66 (10.2%) |
| **MIA TILs score** |  |  |
| 0 | 138 (24.4%) | 337 (51.8%) |
| 1 | 262 (46.4%) | 209 (32.2%) |
| 2 | 126 (22.3%) | 77 (11.8%) |
| 3 | 39 (6.9%) | 27 (4.2%) |
| **Stromal TIL score (continuous)** |  |  |
| Median, % [IQI] | 10 [5-30] | 5 [0-20] |
| Not enough tumor to be assessed | 23 | 93 |
| **Stromal TIL score (categorical)** |  |  |
| 0% | 113 (20.8%) | 240 (43.1%) |
| 5-10% | 193 (35.6%) | 148 (26.6%) |
| 15-100% | 236 (43.5%) | 169 (30.3%) |
| Not enough tumor to be assessed | 23 | 93 |

### TILs in primary melanoma specimens and association with response and survival

In the group of patients with primary specimens, 24.4% of patients had absent TILs, while 60.7% had non-brisk and 14.9% had brisk TILs. Regarding the MIA score, 46.4% had a score of 1, 22.3% score of 2 and 6.9% score 3. The median of the stromal score was 10% (IQI 5-30%). None of the TIL scores demonstrated a significant association with response (p=0.55, p=0.36, and p=0.44 respectively, supplementary figure 1). Also, no significant association between any TIL score and survival was found in this group (supplementary table 6).

### TILs in metastatic melanoma specimens and association with response

In the group of patients with metastatic specimens available, 10.2% had brisk TILs, and 51.8% had no TILs. A visual representation of brisk, non-brisk and absent TILs is shown in figure 2. Patients who responded to ICI had significantly more TILs compared to patients who did not respond, which was reflected in the Clark, MIA and stromal TIL scores (all p<0.001, figure 3). Furthermore, when categorized on TIL pattern the ORR increased from 46% in patients with absent TILs to 59% and 74% in patients with non-brisk and brisk TILs, respectively. Likewise, patients with MIA score 3 had a higher ORR compared to MIA score 0 (78% vs. 46%, respectively).

**Figure 2A-C.**
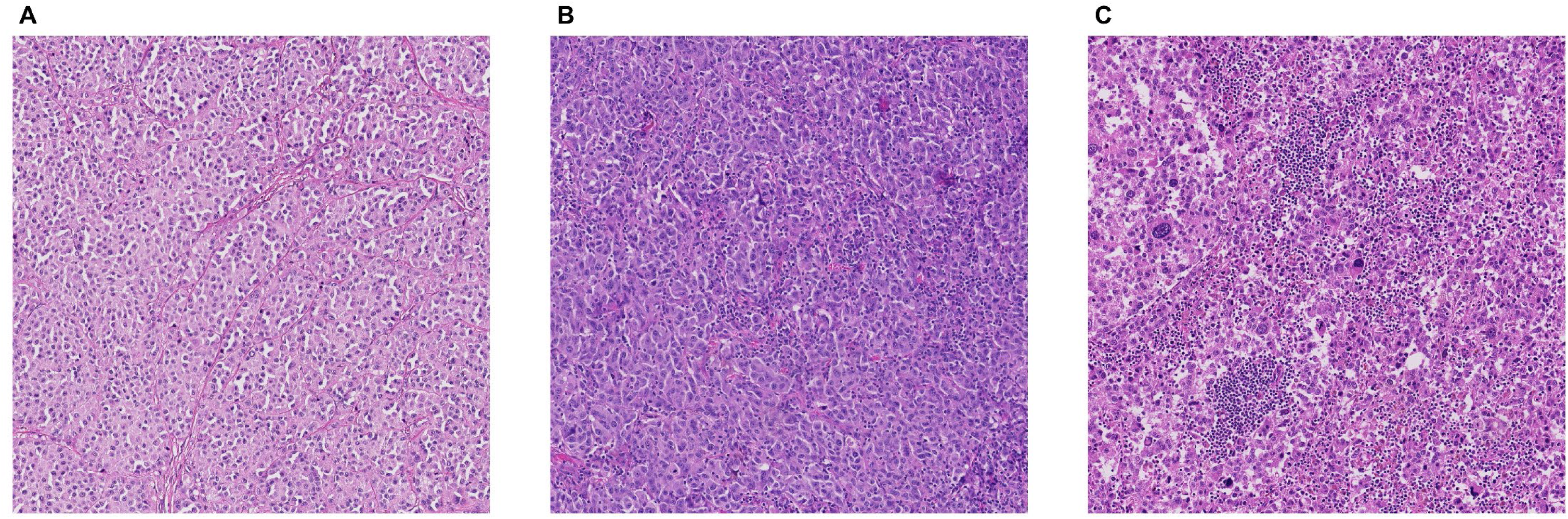
Visual representation of TIL score in pre-treatment metastatic sample. Absent TILs in cutaneous metastasis, (B) non-brisk TILs in lymph node metastasis, (C) brisk TILs in lymph node metastasis.

**Figure 3A-C.**
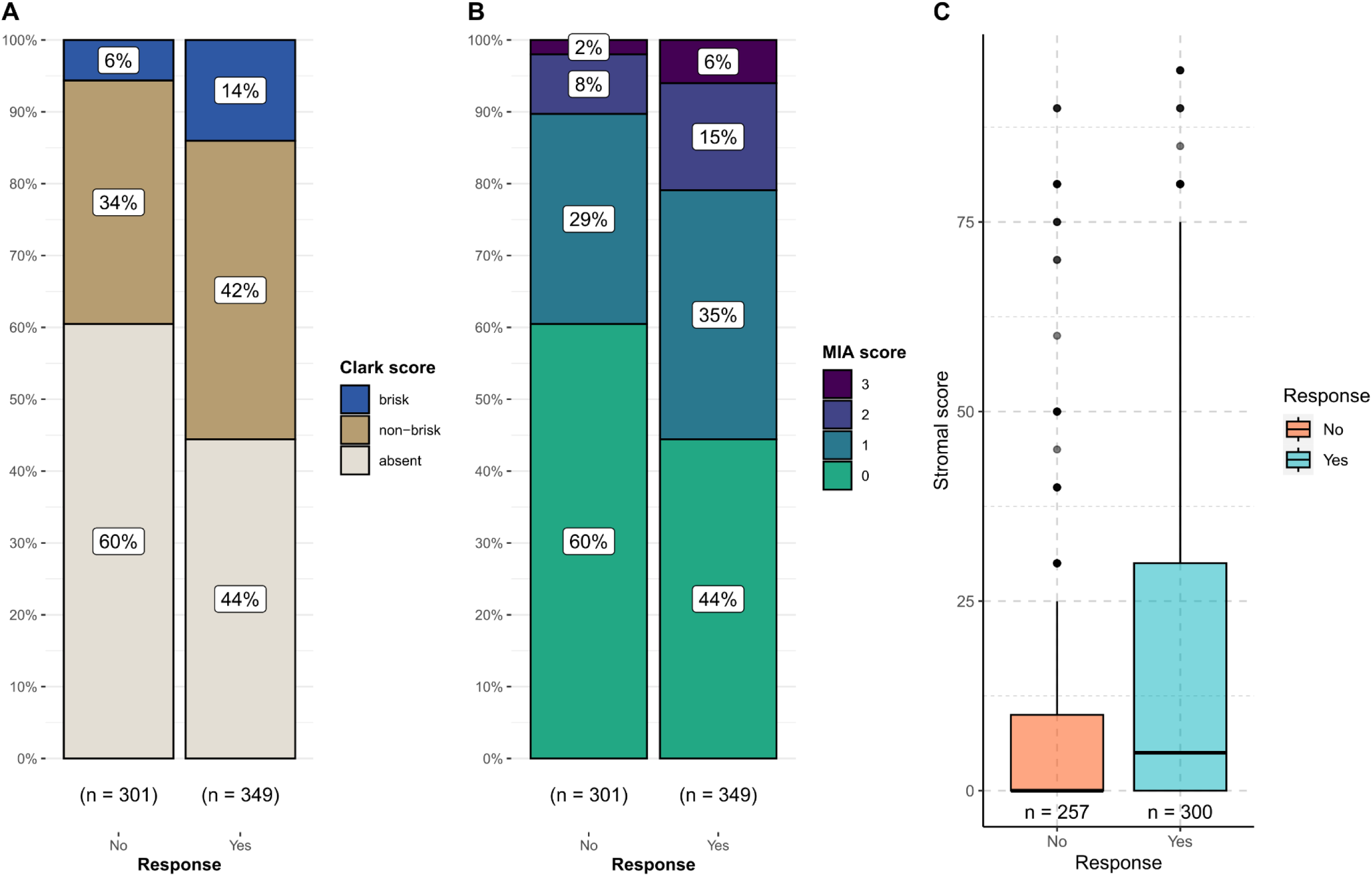
Comparison of response to ICI in patients with available metastatic melanoma specimen categorized by TILs score. Bar charts and boxplot differentiating between response (‘yes’) and no response (‘no’). (A) Patients categorized by Clark score (p<0.001), (B) patients categorized by MIA score (p<0.001) and (C) Stromal score plotted on y-axis (p<0.001). The numbers below the plots reflect the number of patients in the corresponding groups. In (C), there are 93 less patients because of unavailability of the stromal score, because of the absence of tumoral stroma.

In univariable analysis with the Clark score, both non-brisk TILs and brisk-TILs showed a significant association with ICI response compared to absent TILs (OR 1.67 [95%CI 1.20-2.33] and 3.38 [95%CI 1.91-6.27], respectively). Having a MIA score of 1, 2 or 3 was also significantly associated with response to ICI, compared to score 0 (OR 1.61 [95% CI 1.14-2.29] for MIA score 1, OR 2.44 [95% CI 1.46-4.17] for MIA score 2, and OR 4.11 [95% CI1.71-11.43] for MIA score 3). Regarding the stromal score as a continuous variable, there was also a significant association with response (OR 1.02 [95%CI 1.01-1.02]). When using the stromal score as a categorial variable, a stromal score of 15% or higher was significantly associated with a higher chance of response (OR 2.35 [95%CI 1.56-3.54]). After adjusting for age, sex, stage of disease, WHO performance score, level of LDH, presence of BRAF V600 mutation, symptomatic brain metastasis, and type of therapy in multivariable analysis, the association remained significant across all three of the scoring systems. Table 2 shows the results of univariable and multivariable analyses (for the full logistic regression analysis, see supplementary tables 7-9).

**Table 2.** Univariable and multivariable logistic regression analysis of TIL scoring systems with response to ICI for 645 ICI-treated advanced cutaneous melanoma patients, of whom the pre-treatment metastatic specimen was available for analysis. In the multivariable analyses, type of therapy, age, sex, stage of disease, WHO performance score, level of LDH, presence of symptomatic brain metastases, and presence of BRAF V600 mutation were taken into account. For each TIL score, a separate analysis was performed. P-values for median survival times were calculated using logrank tests. Odds ratios (OR) with 95% confidence intervals (CI), and median PFS and OS in months with 95% CI’s are shown.

| TILs scoring system |  | Univariable analysis |  |  | Multivariable analysis |  |  | Objective response rate |
| --- | --- | --- | --- | --- | --- | --- | --- | --- |
|  |  | OR | 95% CI | p-value | OR | 95% CI | p-value | % |
| Clark score | *Absent* | REF |  |  | REF |  |  | 46 |
|  | *Non-brisk* | 1.67 | 1.20-2.33 | 0.002 | 1.56 | 1.06-2.29 | 0.024 | 59 |
|  | *Brisk* | 3.38 | 1.91-6.27 | <0.001 | 3.28 | 1.72-6.56 | <0.001 | 74 |
| MIA score | *0* | REF |  |  | REF |  |  | 46 |
|  | *1* | 1.61 | 1.14-2.29 | 0.007 | 1.52 | 1.02-2.29 | 0.042 | 57 |
|  | *2* | 2.44 | 1.46-4.17 | <0.001 | 2.29 | 1.27-4.20 | 0.006 | 69 |
|  | *3* | 4.11 | 1.71-11.43 | 0.003 | 3.62 | 1.42-10.6 | 0.011 | 78 |
| Stromal score | *Continuous* | 1.02 | 1.01-1.02 | <0.001 | 1.02 | 1.01-1.03 | <0.001 |  |
| Stromal score | *0%* | REF |  |  | REF |  |  | 46 |
|  | *5-10%* | 1.23 | 0.81-1.85 | 0.329 | 1.38 | 0.86-2.22 | 0.181 | 51 |
|  | *15-100%* | 2.35 | 1.56-3.54 | <0.001 | 2.30 | 1.43-3.72 | <0.001 | 66 |

### TIL scores on metastatic specimens and survival

Patients with brisk or non-brisk TILs in their metastatic specimens had a longer median PFS compared to patients with absent TILs (19.4 and 9.2 months vs. 6.5 months, respectively; p=0.009). Moreover, these groups had a longer median OS (40.9 and 49.5 months vs. 21.3 months, p=0.007). Upon stratification based on the MIA score, patients with a score of 2 or 3 had longer median PFS compared to patients with a score of 0 (18.6 and 17.6 months vs. 6.5 months, respectively; p=0.013). Similarly, patients with a MIA score of 2 or 3 had a longer median OS compared to patients with a score of 0 (40.9 months and median not reached vs. 21.3 months, respectively; p=0.010). Patients with higher stromal TIL scores experienced longer survival than those without TILs. When stratifying patients based on categories of the stromal score, those who had 15% or more stromal TILs had longer median PFS (13.2 months vs. 7.4 months, p=0.048) and OS (49.5 months vs. 19.5 months; p=0.003) compared to 0%.

Kaplan-Meier curves for PFS and OS for the Clark score are depicted in figure 4, and median survival times for the TIL scores are shown in table 2. Supplementary figure 2 show Kaplan-Meier curves for PFS and OS for the other two TIL scores.

**Figure 4.**
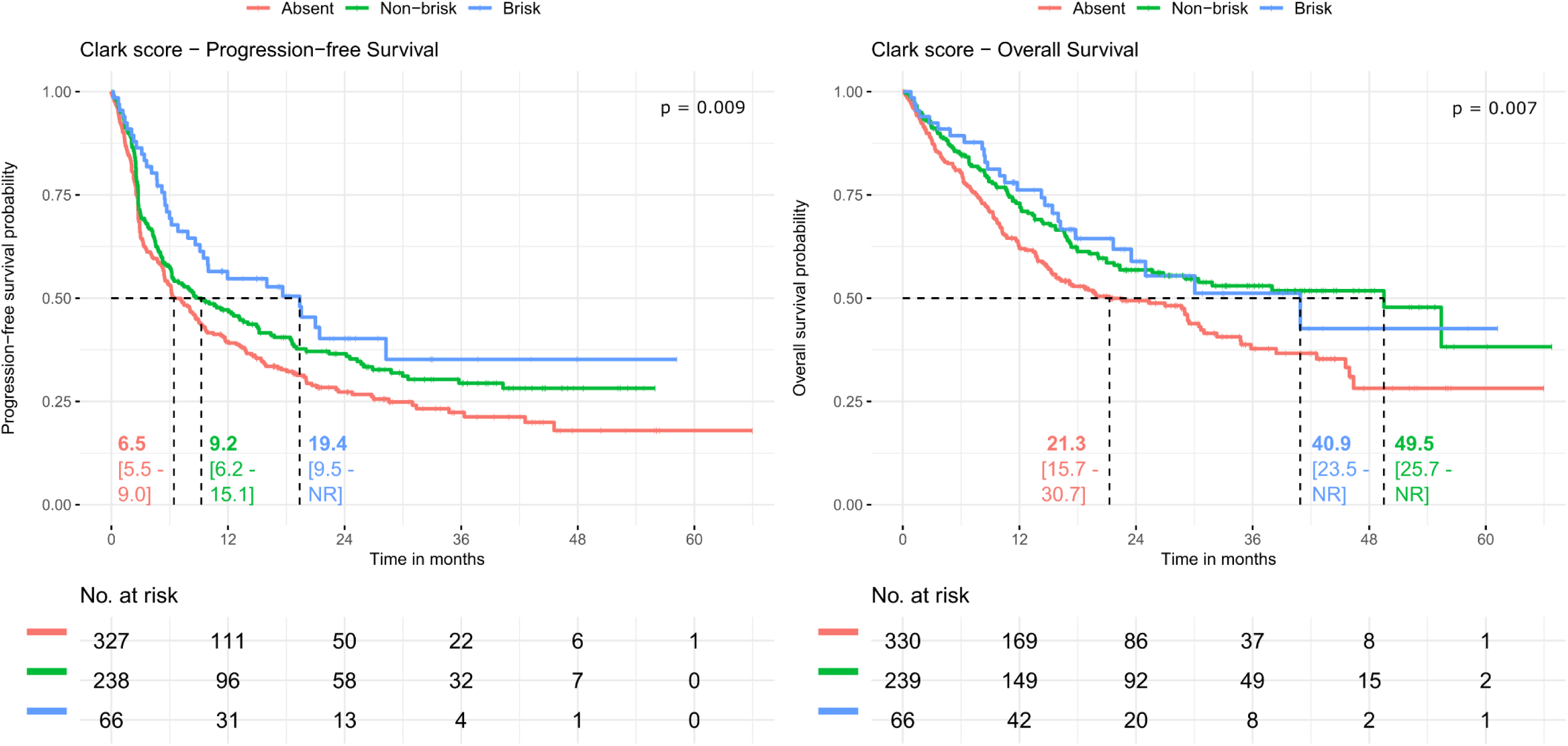
Kaplan Meier curves for progression-free survival (PFS) and overall survival (OS) for the Clark score in patients with a metastatic melanoma specimen with median survival times and corresponding 95% confidence interval. P-values for median survival times were calculated using logrank tests. In these curves, 19 patients were not included in the PFS analysis, and 15 patients were not included in the OS analysis because of missing survival outcomes. Abbreviations: NR, not reached.

In multivariable Cox regression analysis adjusting for known clinical predictors, having more TILs was significantly associated with a prolonged PFS regarding both the Clark score and the MIA score. Additionaly, for each TIL score, a trend towards a prolonged OS was observed in patients who had more TILs. Results for the multivariable Cox proportional hazard regression analysis are shown in supplementary table 10-12.

### Comparison of TILs in primary melanoma and matched metastatic specimens

For 341 patients, both primary and pre-treatment metastatic specimens were available. Out of the patients with non-brisk or brisk TILs in their primary specimen (n=258), 47.3% had an absent score (n=122) in their metastatic specimen. When evaluating primary and metastasic specimens across these patients, there was a significant decrease in the presence of the TILs in the metastasis using the Clark score (supplementary figure 4, p<0.001). This significant difference was also present when using the two other TIL score methods (supplementary figure 4 and table 13).

### Interobserver variability

The degree of interobserver agreement was tested in 50 primary melanoma specimens and 50 metastastic specimens. Interobserver agreement was substantial for the Clark score in both primary specimens (κ = 0.75) and pre-treatment metastatic specimens (κ = 0.78). For the MIA score, the degree of interobserver agreement was moderate (κ = 0.66 for primary melanomas and κ = 0.63 for pre-treatment metastatic specimens). Regarding the stromal score, the intra-class correlation coefficient was substantial for primary melanoma specimens (0.85) and metastatic melanoma specimens (0.73).

## Discussion

In the present study, we showed that in patients with metastatic melanoma, the presence of TILs in pre-treatment metastatic specimens was associated with a better response to ICI, and with longer PFS and OS. These observations held true for all three TIL scoring systems, with associations being independent of known clinical predictors. No relationship between TIL score of the primary melanoma and response to ICI or survival in the metastatic setting was found. The TIL scores were significantly different between matched primary melanoma and metastatic specimens, with a notable decrease in the presence of TILs in metastatic specimens.

Earlier smaller studies have investigated the relationship between TILs and response to ICI, which encompassed mostly anti-PD-1 treatment regimens. Stephens *et al* found that the absence of TILs in primary melanomas was associated with a greater risk for progressive disease after ICI for metastatic disease.^13^ However, their study involved a cohort of only 142 patients and the TIL assessment was based on information extracted from pathology reports rather than being reassessed by the authors themselves. In our larger cohort, when assessing TILs in primary melanoma specimens, we found no relationship between primary melanoma TILs and response to ICI. Our findings are in line with the study of Straker *et al.,* who also found that non-brisk and brisk TILs in primary specimens on H&E slides were not associated with an improved PFS in 114 ICI advanced melanoma patients treated with ICI.^14^

In metastatic melanoma, several smaller studies found an association between CD8^+^ TILs and a response to anti-PD1, anti-CTLA4 and combination therapy.^18,19^ However, to determine CD8^+^ TILs, immunohistochemistry methods are required. On H&E slides, two studies that looked specifically into TILs on metastatic specimens and response to ICI found conflicting results, which might be due to the both studies having a small cohort of less than 20 patients. ^20,21^ More recently, machine learning algorithms were used to recognize eTILs in H&E slides of metastatic melanoma. The presence and amount of eTILs in metastatic specimens was associated with response to anti-PD1 and prolonged OS and PFS.^22^ In neo-adjuvant anti-PD-1 treated patients, brisk TILs in the post-treatment metastasis have been associated with prolonged disease free survival and an increase in pathological response.^23^

Regarding other diseases, such as renal cell carcinoma, the presence of TILs on H&E slides of pre-treatment metastatic specimens has also been linked to response to ICI.^24^ Our research, the largest study to date, shows a clear association between TILs on H&E slides of pre-treatment metastasis and ICI response. Furthermore this study is the first to show that this association is independent of known clinical predictive factors.

Based on our results, TILs in primary melanoma specimens do not predict ICI treatment response or survival in patients with advanced melanoma. As an explanation, it can be hypothesized that a primary melanoma with an appropriate immune response is less likely to metastasize. Another study comparing TIL density in primary melanoma and metastases also found substantial differences in TILs between primary melanomas and metastases, but a high similarity in TILs between different metastatic lesions within individual patients.^25^ This discrepancy is reflected in our cohort by a lower percentage of specimens with TILs present in our metastatic cohort. The presence of TILs in metastatic specimens likely is a more adequate reflection of the situation at the start of therapy, which explains why the presence of pre-treatment metastatic TILs is associated with ICI-response.

We are the first to compare three different TILs scoring systems on H&E-stained specimens in ICI-treated patients with advanced melanoma. The Clark, MIA and stromal TILs scoring system were all associated with better response to ICI and better survival. Interobserver agreement was highest in the Clark score and in line with previous literature.^26^ In our experience, the Clark scoring system that differentiates between absent, non-brisk and brisk TILs is best reproducible, and is most widely used. Therefore, we would suggest using the Clark TILs score when assessing TILs on H&E-stained slides in metastatic melanoma specimens.

The strengths of our work are the large size of our cohort, the multicenter design and the adjustment for known clinical predictors, which shows that the association is independent. As described above, this is the largest study describing the presence of TILs in both primary melanoma and pre-treatment metastatic specimen, which adds to the weight of our presented conclusions. Furthermore, the dataset includes patients from ten academic and non-academic hospitals and is thus representative for a general advanced melanoma population undergoing ICI treatment. A potential limitation is the exclusion of patients due to unavailability of either the primary melanoma or the pre-treatment metastatic specimen. However, we do not think this led to bias, because the baseline characteristics of included and excluded patients were comparable. The substantial interobserver agreement is still subjective to differences due to interobserver variability which may form a limition in the generalizability of the results. Future studies should be directed at AI guided TIL detection to eliminate the differences caused by interobser variability.

Concluding, in advanced melanoma patients, TILs in H&E-stained pre-treatment metastatic specimens are associated with an increased response to ICI, which also translated into prolonged survival. This association remains after adjustment for known clinical predictors of ICI response in melanoma. Pathologists may therefore consider including the Clark TIL score of pre-treatment metastatic specimens in their pathology report. In future research, these scores could be incorporated in multimodal prediction models to better predict which patient will respond to ICI.

## Data Availability

Due to confidentiality agreements, clinical and imaging data cannot be made available.

## Author Contributions

Concept and design: van Duin, Schuiveling, Blokx, van Diest, Suijkerbuijk

Acquisition, analysis, or interpretation of data: van Duin, Schuiveling, ter Maat, Blokx, van Diest, Suijkerbuijk

Drafting of the manuscript: van Duin, Schuiveling, ter Maat, van Amsterdam, van den Berkmortel, Boers-Sonderen, Hospers, Labots, de Groot, Kapiteijn, Piersma, Vreugdenhil, Westgeest, Schrader, Veta, Blokx, van Diest, Suijkerbuijk

Statistical analysis: van Duin, Schuiveling

Administrative, technical, or material support: van den Berkmorten, Boers-Sonderen, Hospers, Labots, de Groot, Kapiteijn, Piersma, Vreugdenhil, Westgeest, Suijkerbuijk Supervision: Blokx, van Diest, Suijkerbuijk

## References

1. Wolchok, J. D. et al. Long-Term Outcomes With Nivolumab Plus Ipilimumab or Nivolumab Alone Versus Ipilimumab in Patients With Advanced Melanoma. JCO 40, 127–137 (2022).

2. van Not, O. J., et al. BRAF and NRAS Mutation Status and Response to Checkpoint Inhibition in Advanced Melanoma. JCO Precis Oncol 6, e2200018 (2022).

3. Postow, M. A., Sidlow, R. & Hellmann, M. D. Immune-Related Adverse Events Associated with Immune Checkpoint Blockade. N Engl J Med 378, 158–168 (2018).

4. Franken, M. G. et al. Trends in survival and costs in metastatic melanoma in the era of novel targeted and immunotherapeutic drugs. ESMO Open 6, 100320 (2021).

5. Clark, W. H., From, L., Bernardino, E. A. & Mihm, M. C. The histogenesis and biologic behavior of primary human malignant melanomas of the skin. Cancer Res 29, 705–727 (1969).

6. Fu, Q. et al. Prognostic value of tumor-infiltrating lymphocytes in melanoma: a systematic review and meta-analysis. Oncoimmunology 8, 1593806 (2019).

7. Brummel, K., Eerkens, A. L., de Bruyn, M. & Nijman, H. W. Tumour-infiltrating lymphocytes: from prognosis to treatment selection. Br J Cancer 128, 451–458 (2023).

8. Clark, W. H., Jr., et al. Model Predicting Survival in Stage I Melanoma Based on Tumor Progression. JNCI: Journal of the National Cancer Institute 81, 1893–1904 (1989).

9. Azimi, F. et al. Tumor-Infiltrating Lymphocyte Grade Is an Independent Predictor of Sentinel Lymph Node Status and Survival in Patients With Cutaneous Melanoma. JCO 30, 2678–2683 (2012).

10. Hendry, S. et al. Assessing tumor infiltrating lymphocytes in solid tumors: a practical review for pathologists and proposal for a standardized method from the International Immuno-Oncology Biomarkers Working Group. Adv Anat Pathol 24, 311–335 (2017).

11. Fairfax, B. P. et al. Peripheral CD8+ T cell characteristics associated with durable responses to immune checkpoint blockade in patients with metastatic melanoma. Nat Med 26, 193–199 (2020).

12. Lee, J. S. & Ruppin, E. Multiomics Prediction of Response Rates to Therapies to Inhibit Programmed Cell Death 1 and Programmed Cell Death 1 Ligand 1. JAMA Oncology 5, 1614–1618 (2019).

13. Stephens, M. R. et al. Association Between Metastatic Melanoma Response to Checkpoint Inhibitor Therapy and Tumor-Infiltrating Lymphocyte Classification on Primary Cutaneous Melanoma Biopsies. JAMA Dermatology 159, 215–216 (2023).

14. Straker, R. J. et al. Prognostic significance of primary tumor infiltrating lymphocytes in a contemporary melanoma cohort. Ann Surg Oncol 29, 5207–5216 (2022).

15. Jochems, A. et al. Dutch Melanoma Treatment Registry: Quality assurance in the care of patients with metastatic melanoma in the Netherlands. Eur J Cancer 72, 156–165 (2017).

16. Gershenwald, J. E., et al. Melanoma staging: Evidence-based changes in the American Joint Committee on Cancer eighth edition cancer staging manual. CA: A Cancer Journal for Clinicians 67, 472–492 (2017).

17. Eisenhauer, E. A. et al. New response evaluation criteria in solid tumours: Revised RECIST guideline (version 1.1). European Journal of Cancer 45, 228–247 (2009).

18. Tumeh, P. C. et al. PD-1 blockade induces responses by inhibiting adaptive immune resistance. Nature 515, 568–571 (2014).

19. Hamid, O. et al. Safety, Clinical Activity, and Biological Correlates of Response in Patients with Metastatic Melanoma: Results from a Phase I Trial of Atezolizumab. Clinical Cancer Research 25, 6061–6072 (2019).

20. Taube, J. M. et al. Association of PD-1, PD-1 Ligands, and Other Features of the Tumor Immune Microenvironment with Response to Anti–PD-1 Therapy. Clinical Cancer Research 20, 5064–5074 (2014).

21. Mastracci, L. et al. Response to ipilimumab therapy in metastatic melanoma patients: potential relevance of CTLA-4+ tumor infiltrating lymphocytes and their in situ localization. Cancer Immunol Immunother 69, 653–662 (2020).

22. Chatziioannou, E. et al. Deep learning-based scoring of tumour-infiltrating lymphocytes is prognostic in primary melanoma and predictive to PD-1 checkpoint inhibition in melanoma metastases. eBioMedicine 93, 104644 (2023).

23. Huang, A. C. et al. A Single Dose of Neoadjuvant PD-1 Blockade Predicts Clinical Outcomes in Resectable Melanoma. Nat Med 25, 454–461 (2019).

24. Deutsch, J. S. et al. Combinatorial biomarker for predicting outcomes to anti-PD-1 therapy in patients with metastatic clear cell renal cell carcinoma. Cell Reports Medicine 4, 100947 (2023).

25. Gorris, M. A. J. et al. Paired primary and metastatic lesions of patients with ipilimumab-treated melanoma: high variation in lymphocyte infiltration and HLA-ABC expression whereas tumor mutational load is similar and correlates with clinical outcome. J Immunother Cancer 10, e004329 (2022).

26. Busam, K. J. et al. Histologic classification of tumor-infiltrating lymphocytes in primary cutaneous malignant melanoma. A study of interobserver agreement. Am J Clin Pathol 115, 856–860 (2001).

