## Supplements for "Baseline tumor-infiltrating lymphocyte patterns and response to immune checkpoint inhibition in metastatic cutaneous melanoma"

**Supplementary material**

Supplementary table 1. TIL scoring system as described by Clark *et al*, further elaborated by Clemente *et al*.[1,2]

| TIL grade | Explanation |
|---|---|
| Absent | No TILs in tumor tissue |
| Non-brisk - localized | Patchy multifocal lymphocytic infiltrate or mildly diffuse, but overall little TILs throughout the tumor |
| Brisk | TILs infiltrate the entire base of the tumor and/or diffusely infiltrate the entire invasive component of the tumor |

Supplementary table 2. TIL scoring system as described by the Melanoma Institute Australia (MIA).[3]

| TIL grade | Explanation |
|---|---|
| 0 | Absent |
| 1 | Mild infiltrate of lymphocytes focally interspersed among the melanoma cells. |
| 2 | Dense multifocal infiltrate of lymphocytes interspersed among the melanoma cells. |
| 3 | Dense infiltrate of lymphocytes illustrated in the figure was present diffusely throughout the tumor. |

Supplementary table 3. Stepwise approach of assessing TILs in the tumoral stroma, as described by Hendry.[4]

| | |
|---|---|
| Step 1 | Define the tumor area. Exclude areas with immune infiltrate outside tumor border. |
| Step 2 | Distinguish between intratumoral TILs and tumoral stromal TIL's |
| Step 3 | Identify the type of inflammatory infiltrate, excluding granulocytes and necrosis. |
| Step 4 | Assess the percentage of TILs. |
| | In our analysis, The percentage can range from 0 to 100%, in increments of 5%. |

Supplementary table 4. Patient characteristics of included (not having a pre-treatment metastatic melanoma specimen available) versus excluded (not having a pre-treatment metastatic melanoma specimen available) patients

| | Excluded (N=647) | Included (N=650) | Total (N=1297) |
|---|---|---|---|
| **Age (years)** | | | |
| Median [IQI] | 69 [58-76] | 66 [57-74] | 68 [57- 75] |
| **Sex** | | | |
| Female | 265 (41.0%) | 228 (35.1%) | 493 (38.0%) |
| Male | 382 (59.0%) | 422 (64.9%) | 804 (62.0%) |
| **WHO Performance status** | | | |
| WHO 0 | 325 (52.9%) | 285 (45.8%) | 610 (49.4%) |
| WHO 1 | 228 (37.1%) | 291 (46.8%) | 519 (42.0%) |
| WHO 2-4 | 61 (9.9%) | 46 (7.4%) | 107 (8.7%) |
| Missing | 33 | 28 | 61 |
| **Stage of disease** | | | |
| Unresectable IIIC | 56 (9.0%) | 50 (8.1%) | 106 (8.5%) |
| M1a | 52 (8.4%) | 44 (7.1%) | 96 (7.7%) |
| M1b | 93 (15.0%) | 81 (13.1%) | 174 (14.0%) |
| M1c | 274 (44.1%) | 276 (44.5%) | 550 (44.3%) |
| M1d - non symptomatic | 86 (13.8%) | 93 (15.0%) | 179 (14.4%) |
| M1d - symptomatic | 61 (9.8%) | 76 (12.3%) | 137 (11.0%) |
| Missing | 25 | 30 | 55 |
| **BRAF V600 Mutation** | | | |
| Wildtype | 408 (70.0%) | 404 (70.3%) | 812 (70.1%) |
| Mutant | 175 (30.0%) | 171 (29.7%) | 346 (29.9%) |
| Missing | 64 | 75 | 139 |
| **LDH levels** | | | |
| Not elevated | 438 (68.4%) | 413 (64.4%) | 851 (66.4%) |
| 1-2x ULN | 152 (23.8%) | 180 (28.1%) | 332 (25.9%) |
| >2x ULN | 50 (7.8%) | 48 (7.5%) | 98 (7.7%) |
| Missing | 7 | 9 | 16 |
| **Type of systemic therapy** | | | |
| Anti-PD1 | 445 (68.8%) | 401 (61.7%) | 846 (65.2%) |
| Ipilimumab & Nivolumab | 202 (31.2%) | 249 (38.3%) | 451 (34.8%) |
| **Response** | | | |
| No | 309 (47.8%) | 301 (46.3%) | 610 (47.0%) |
| Yes | 338 (52.2%) | 349 (53.7%) | 687 (53.0%) |
| **Progression-free survival (months)** | | | |
| Median (95% CI) | 8.3 (6.6-10.8) | 8.6 (6.8-11.0) | 8.5 (7.2-9.8) |
| **Overall survival (months)** | | | |
| Median (95% CI) | 36.5 (27.1-56.6) | 29.4 (23.5-38.4) | 31.8 (27.2-40.1) |

Supplementary table 5. Organ localization of the pre-treatment metastatic samples.

| **Organ localisation** | **Number of patients** |
|---|---|
| Lymphnode | 195 |
| Skin | 134 |
| Soft tissue | 68 |
| Lung | 58 |
| Liver | 39 |
| Brain | 39 |
| Bone | 20 |
| Colon | 8 |
| Stomach | 8 |
| Small intestine | 7 |
| Adrenal gland | 4 |
| Oesophagus | 2 |
| Gallbladder | 2 |
| Pancreas | 1 |
| Urogenital tract | 1 |
| Unknown | 64 |

Supplementary table 6. Multivariable Cox proportional hazard regression for PFS and OS in patients with primary specimen available. In the multivariable analyses, type of therapy, age, sex, stage of disease, WHO performance score, level of LDH, presence of symptomatic brain metastases, and presence of BRAF V600 mutation were taken into account. For each TILs score, a separate analysis was performed. Hazard ratios (HR) and 95% confidence intervals (CI) are shown.

| | Progression-free survival | | | Overall survival | | |
|---|---|---|---|---|---|---|
| **Characteristic** | **HR**[1] | **95% CI**[1] | **p-value** | **HR**[1] | **95% CI**[1] | **p-value** |
| Clark score | | | | | | |
| absent | — | — | | — | — | |
| brisk | 0.84 | 0.64-1.11 | 0.2 | 0.85 | 0.60-1.19 | 0.3 |
| non-brisk | 0.83 | 0.57-1.23 | 0.4 | 0.89 | 0.56-1.43 | 0.6 |
| MIA score | | | | | | |
| 0 | — | — | | — | — | |
| 1 | 0.88 | 0.66-1.17 | 0.4 | 0.87 | 0.61-1.23 | 0.4 |
| 2 | 0.80 | 0.57-1.13 | 0.2 | 0.95 | 0.63-1.43 | 0.8 |
| 3 | 0.73 | 0.45-1.20 | 0.2 | 0.56 | 0.29-1.07 | 0.079 |
| Stromal score | 1.00 | 1.0-1.01 | >0.9 | 1.00 | 0.99-1.01 | 0.9 |

[1] HR = Hazard Ratio, CI = Confidence Interval

Supplementary table 7. Complete multivariable logistic regression analysis of Clark score with response to ICI for patients of whom the pre-treatment metastatic specimen was available for analysis.

| **Characteristic** | **OR**[1] | **95% CI**[1] | **p-value** |
|---|---|---|---|
| Clark TILs score | | | |
| Absent | — | — | |
| Non-brisk | 1.56 | 1.06, 2.29 | 0.024 |
| Brisk | 3.28 | 1.72, 6.56 | <0.001 |
| Age | 1.00 | 0.99, 1.02 | 0.6 |
| Sex | | | |
| Female | — | — | |
| Male | 1.15 | 0.78, 1.69 | 0.5 |
| Stage of disease | | | |
| Unresectable IIIC | — | — | |
| M1a | 3.18 | 1.20, 8.92 | 0.023 |
| M1b | 1.65 | 0.73, 3.74 | 0.2 |
| M1c | 1.09 | 0.54, 2.20 | 0.8 |
| M1d - non symptomatic | 1.21 | 0.54, 2.72 | 0.6 |
| M1d - symptomatic | 0.54 | 0.23, 1.26 | 0.2 |
| WHO performance score | | | |
| 0 | — | — | |
| 1 | 1.11 | 0.75, 1.64 | 0.6 |
| 2-4 | 0.96 | 0.45, 2.05 | >0.9 |
| Level of LDH | | | |
| Not elevated | — | — | |
| Elevated, 1-2x ULN | 0.94 | 0.61, 1.45 | 0.8 |
| Elevated, >2x ULN | 0.35 | 0.16, 0.75 | 0.008 |
| Mutation Status | | | |
| Wildtype | — | — | |
| BRAF V600 E mutation present | 1.09 | 0.73, 1.64 | 0.7 |
| Type of therapy | | | |
| Anti-PD1 | — | — | |
| Ipilimumab & Nivolumab | 1.29 | 0.84, 1.99 | 0.2 |

[1] OR = Odds Ratio, CI = Confidence Interval

Supplementary table 8. Complete multivariable logistic regression analysis of MIA score with response to ICI for patients of whom the pre-treatment metastatic specimen was available for analysis.

| **Characteristic** | **OR**[1] | **95% CI**[1] | **p-value** |
|---|---|---|---|
| MIA TILs score | | | |
| 0 | — | — | |
| 1 | 1.52 | 1.02, 2.29 | 0.042 |
| 2 | 2.29 | 1.27, 4.20 | 0.006 |
| 3 | 3.62 | 1.42, 10.6 | 0.011 |
| Age | 1.00 | 0.99, 1.02 | 0.6 |
| Sex | | | |
| Female | — | — | |
| Male | 1.14 | 0.77, 1.68 | 0.5 |
| Stage of disease | | | |
| Unresectable IIIC | — | — | |
| M1a | 3.20 | 1.21, 8.96 | 0.022 |
| M1b | 1.60 | 0.71, 3.64 | 0.3 |
| M1c | 1.09 | 0.54, 2.19 | 0.8 |
| M1d - non symptomatic | 1.20 | 0.54, 2.69 | 0.7 |
| M1d - symptomatic | 0.53 | 0.23, 1.24 | 0.15 |
| WHO performance score | | | |
| 0 | — | — | |
| 1 | 1.09 | 0.74, 1.61 | 0.7 |
| 2-4 | 0.95 | 0.45, 2.03 | >0.9 |
| Level of LDH | | | |
| Not elevated | — | — | |
| Elevated, 1-2x ULN | 0.93 | 0.61, 1.44 | 0.7 |
| Elevated, >2x ULN | 0.36 | 0.16, 0.76 | 0.010 |
| Mutation Status | | | |
| Wildtype | — | — | |
| BRAF V600 E mutation present | 1.06 | 0.71, 1.59 | 0.8 |
| Type of therapy | | | |
| Anti-PD1 | — | — | |
| Ipilimumab & Nivolumab | 1.28 | 0.83, 1.98 | 0.3 |

[1] OR = Odds Ratio, CI = Confidence Interval

Supplementary table 9. Complete multivariable logistic regression analysis of the stromal score (either as a continuous variable or a categorical variable in the model) with response to ICI for patients of whom the pre-treatment metastatic specimen was available for analysis.

| **Characteristic** | **OR**[1] | **95% CI**[1] | **p-value** |
|---|---|---|---|
| Stromal score (continuous) | 1.02 | 1.01, 1.03 | <0.001 |
| Stromal score (categorical) | | | |
| 0% | — | — | |
| 5-10% | 1.38 | 0.86, 2.22 | 0.2 |
| 15-100% | 2.30 | 1.43, 3.72 | <0.001 |
| Age | 1.01 | 0.99, 1.02 | 0.5 |
| Sex | | | |
| Female | — | — | |
| Male | 0.99 | 0.65, 1.50 | >0.9 |
| Stage of disease | | | |
| Irresectable IIIC | — | — | |
| M1a | 3.12 | 1.11, 9.24 | 0.035 |
| M1b | 1.77 | 0.73, 4.37 | 0.2 |
| M1c | 1.35 | 0.62, 2.92 | 0.4 |
| M1d - non symptomatic | 1.29 | 0.54, 3.12 | 0.6 |
| M1d - symptomatic | 0.57 | 0.22, 1.45 | 0.2 |
| WHO performance score | | | |
| 0 | — | — | |
| 1 | 1.02 | 0.67, 1.54 | >0.9 |
| 2-4 | 0.96 | 0.42, 2.19 | >0.9 |
| Level of LDH | | | |
| Not elevated | — | — | |
| Elevated 1-2x ULN | 1.00 | 0.62, 1.59 | >0.9 |
| Elevated >2x ULN | 0.30 | 0.13, 0.69 | 0.005 |
| Mutation Status | | | |
| Wildtype | — | — | |
| BRAF V600 E mutation present | 0.86 | 0.55, 1.33 | 0.5 |
| Type of therapy | | | |
| Anti-PD1 | — | — | |
| Ipilimumab & Nivolumab | 1.45 | 0.91, 2.35 | 0.12 |

[1] OR = Odds Ratio, CI = Confidence Interval

Supplementary table 10. Complete cox proportional hazard regression analysis with Clark TILs score for PFS and OS for 645 ICI-treated advanced cutaneous melanoma patients, of whom the pre-treatment metastatic specimen was available for analysis.

| **Characteristic** | PFS | | | OS | | |
|---|---|---|---|---|---|---|
| | **HR**[1] | **95% CI**[1] | **p-value** | **HR**[1] | **95% CI**[1] | **p-value** |
| Clark score | | | | | | |
| absent | — | — | | — | — | |
| non-brisk | 0.86 | 0.68, 1.08 | 0.2 | 0.78 | 0.59, 1.03 | 0.081 |
| brisk | 0.56 | 0.38, 0.84 | 0.005 | 0.74 | 0.47, 1.17 | 0.2 |
| Age | 1.00 | 0.99, 1.01 | 0.4 | 1.01 | 1.00, 1.03 | 0.023 |
| Sex | | | | | | |
| Female | — | — | | — | — | |
| Male | 1.02 | 0.81, 1.27 | 0.9 | 0.99 | 0.75, 1.30 | >0.9 |
| Stage of disease | | | | | | |
| Unresectable IIIC | — | — | | — | — | |
| M1a | 1.03 | 0.58, 1.82 | >0.9 | 0.90 | 0.40, 2.02 | 0.8 |
| M1b | 0.95 | 0.57, 1.60 | 0.9 | 1.51 | 0.77, 2.95 | 0.2 |
| M1c | 1.15 | 0.73, 1.82 | 0.5 | 1.51 | 0.82, 2.78 | 0.2 |
| M1d - non symptomatic | 1.46 | 0.88, 2.43 | 0.15 | 2.05 | 1.05, 4.02 | 0.037 |
| M1d - symptomatic | 2.00 | 1.19, 3.34 | 0.009 | 3.54 | 1.82, 6.86 | <0.001 |
| WHO performance score | | | | | | |
| 0 | — | — | | — | — | |
| 1 | 0.94 | 0.74, 1.18 | 0.6 | 0.81 | 0.61, 1.07 | 0.14 |
| 2-4 | 1.60 | 1.07, 2.39 | 0.023 | 1.86 | 1.19, 2.90 | 0.007 |
| Level of LDH | | | | | | |
| Normal | — | — | | — | — | |
| Elevated 1-2x ULN | 1.22 | 0.95, 1.57 | 0.13 | 1.19 | 0.88, 1.61 | 0.3 |
| Elevated >2x ULN | 2.15 | 1.44, 3.21 | <0.001 | 2.54 | 1.60, 4.03 | <0.001 |
| Mutation status | | | | | | |
| Wildtype | — | — | | — | — | |
| BRAF V600 E mutation present | 1.04 | 0.82, 1.32 | 0.7 | 0.82 | 0.61, 1.10 | 0.2 |
| Type of therapy | | | | | | |
| Anti-PD1 | — | — | | — | — | |
| Ipilimumab & Nivolumab | 0.77 | 0.59, 1.00 | 0.054 | 0.81 | 0.59, 1.12 | 0.2 |

[1] HR = Hazard Ratio, CI = Confidence Interval

Supplementary table 11. Complete cox proportional hazard regression analysis with MIA TILs score for PFS and OS for patients of whom the pre-treatment metastatic specimen was available for analysis.

| | PFS | | | OS | | |
|---|---|---|---|---|---|---|
| **Characteristic** | **HR**[1] | **95% CI**[1] | **p-value** | **HR**[1] | **95% CI**[1] | **p-value** |
| MIA TILs score | | | | | | |
| 0 | — | — | | — | — | |
| 1 | 0.90 | 0.70, 1.14 | 0.4 | 0.80 | 0.60, 1.08 | 0.14 |
| 2 | 0.63 | 0.44, 0.90 | 0.011 | 0.72 | 0.46, 1.13 | 0.2 |
| 3 | 0.56 | 0.31, 1.01 | 0.055 | 0.66 | 0.33, 1.31 | 0.2 |
| Age | 1.00 | 0.99, 1.01 | 0.4 | 1.01 | 1.00, 1.03 | 0.024 |
| Sex | | | | | | |
| Female | — | — | | — | — | |
| Male | 1.02 | 0.81, 1.28 | 0.9 | 0.99 | 0.75, 1.31 | >0.9 |
| Stage of disease | | | | | | |
| Unresectable IIIC | — | — | | — | — | |
| M1a | 1.02 | 0.57, 1.81 | >0.9 | 0.89 | 0.39, 2.00 | 0.8 |
| M1b | 0.97 | 0.58, 1.63 | >0.9 | 1.49 | 0.76, 2.92 | 0.2 |
| M1c | 1.15 | 0.73, 1.82 | 0.5 | 1.49 | 0.81, 2.74 | 0.2 |
| M1d - non symptomatic | 1.45 | 0.87, 2.42 | 0.2 | 2.03 | 1.03, 3.98 | 0.040 |
| M1d - symptomatic | 2.01 | 1.20, 3.37 | 0.008 | 3.46 | 1.79, 6.72 | <0.001 |
| WHO performance score | | | | | | |
| 0 | — | — | | — | — | |
| 1 | 0.95 | 0.75, 1.19 | 0.6 | 0.81 | 0.61, 1.07 | 0.13 |
| 2-4 | 1.59 | 1.07, 2.38 | 0.023 | 1.86 | 1.19, 2.91 | 0.006 |
| Level of LDH | | | | | | |
| Not elevated | — | — | | — | — | |
| Elevated 1-2x ULN | 1.21 | 0.94, 1.56 | 0.13 | 1.19 | 0.88, 1.61 | 0.3 |
| Elevated >2x ULN | 2.12 | 1.42, 3.15 | <0.001 | 2.55 | 1.61, 4.04 | <0.001 |
| Mutation Status | | | | | | |
| Wildtype | — | — | | — | — | |
| BRAF V600E mutation present | 1.07 | 0.84, 1.35 | 0.6 | 0.82 | 0.61, 1.10 | 0.2 |
| Type of therapy | | | | | | |
| Anti-PD1 | — | — | | — | — | |
| Ipilimumab & Nivolumab | 0.77 | 0.59, 1.01 | 0.055 | 0.81 | 0.59, 1.12 | 0.2 |

[1] HR = Hazard Ratio, CI = Confidence Interval

Supplementary table 12. Complete cox proportional hazard regression analysis with the stromal score (either as a continuous variable or a categorical variable in the model) for PFS and OS for patients of whom the pre-treatment metastatic specimen was available for analysis.

| | PFS | | | OS | | |
|---|---|---|---|---|---|---|
| **Characteristic** | **HR**[1] | **95% CI**[1] | **p-value** | **HR**[1] | **95% CI**[1] | **p-value** |
| Stromal score (continuous) | 0.99 | 0.99, 1.00 | 0.021 | 0.99 | 0.99, 1.00 | 0.037 |
| Stromal score (categorical) | | | | | | |
| 0% | — | — | | — | — | |
| 5-10% | 1.04 | 0.78, 1.37 | 0.8 | 0.86 | 0.61, 1.21 | 0.4 |
| 15-100% | 0.78 | 0.59, 1.04 | 0.091 | 0.68 | 0.48, 0.97 | 0.032 |
| Age | 1.00 | 0.99, 1.01 | 0.8 | 1.01 | 1.0, 1.02 | 0.2 |
| Sex | | | | | | |
| Female | — | — | | — | — | |
| Male | 1.04 | 0.81, 1.33 | 0.8 | 0.94 | 0.70, 1.26 | 0.7 |
| Stage of disease | | | | | | |
| Irresectable IIIC | — | — | | — | — | |
| M1a | 1.13 | 0.61, 2.09 | 0.7 | 1.13 | 0.48, 2.65 | 0.8 |
| M1b | 0.89 | 0.50, 1.58 | 0.7 | 1.52 | 0.73, 3.18 | 0.3 |
| M1c | 1.19 | 0.73, 1.94 | 0.5 | 1.61 | 0.83, 3.13 | 0.2 |
| M1d - non symptomatic | 1.54 | 0.89, 2.66 | 0.12 | 2.19 | 1.05, 4.57 | 0.036 |
| M1d - symptomatic | 1.97 | 1.13, 3.45 | 0.018 | 3.74 | 1.83, 7.66 | <0.001 |
| WHO performace score | | | | | | |
| 0 | — | — | | — | — | |
| 1 | 0.99 | 0.77, 1.27 | >0.9 | 0.86 | 0.63, 1.18 | 0.4 |
| 2-4 | 1.56 | 1.00, 2.44 | 0.048 | 2.08 | 1.28, 3.36 | 0.003 |
| Level of LDH | | | | | | |
| Not elevated | — | — | | — | — | |
| Elevated 1-2x ULN | 1.16 | 0.88, 1.54 | 0.3 | 1.21 | 0.87, 1.68 | 0.3 |
| Elevated >2x ULN | 1.85 | 1.20, 2.85 | 0.006 | 2.23 | 1.34, 3.70 | 0.002 |
| Mutation Status | | | | | | |
| Wildtype | — | — | | — | — | |
| BRAF V600E mutation present | 1.15 | 0.89, 1.49 | 0.3 | 0.79 | 0.57, 1.09 | 0.15 |
| Type of therapy | | | | | | |
| Anti-PD1 | — | — | | — | — | |
| Ipilimumab & Nivolumab | 0.79 | 0.59, 1.05 | 0.10 | 0.80 | 0.57, 1.13 | 0.2 |

[1] HR = Hazard Ratio, CI = Confidence Interval

Supplementary table 13

Crosstable of stromal scores in matched primary and pre-treatment metastatic specimens in 348 patients with advanced cutaneous melanoma treated with ICI. A significant shift in TIL score across the primary and metastatic specimens was found ($p < 0.001$).

| **Salgado stromal score** | **Primary specimen** | **Metastatic specimen** |
|---|---|---|
| **Median [IQI]** | 10 [5 -30] | 0 [0 - 15] |

ICC [intraclass correlation] = 0.17 [95% CI 0.06 – 0.29]

Supplementary figure 1. Comparison of response to ICI in patients with available primary melanoma specimen categorized by TIL score. Bar charts and boxplot differentiating between response ('yes') and no response ('no'). (A) Patients categorized by Clark score (p=0.55), (B) patients categorized by MIA score (p=0.36) and (C) Stromal score plotted on y-axis (p=0.44). The numbers below the plots reflect the number of patients in the corresponding groups.

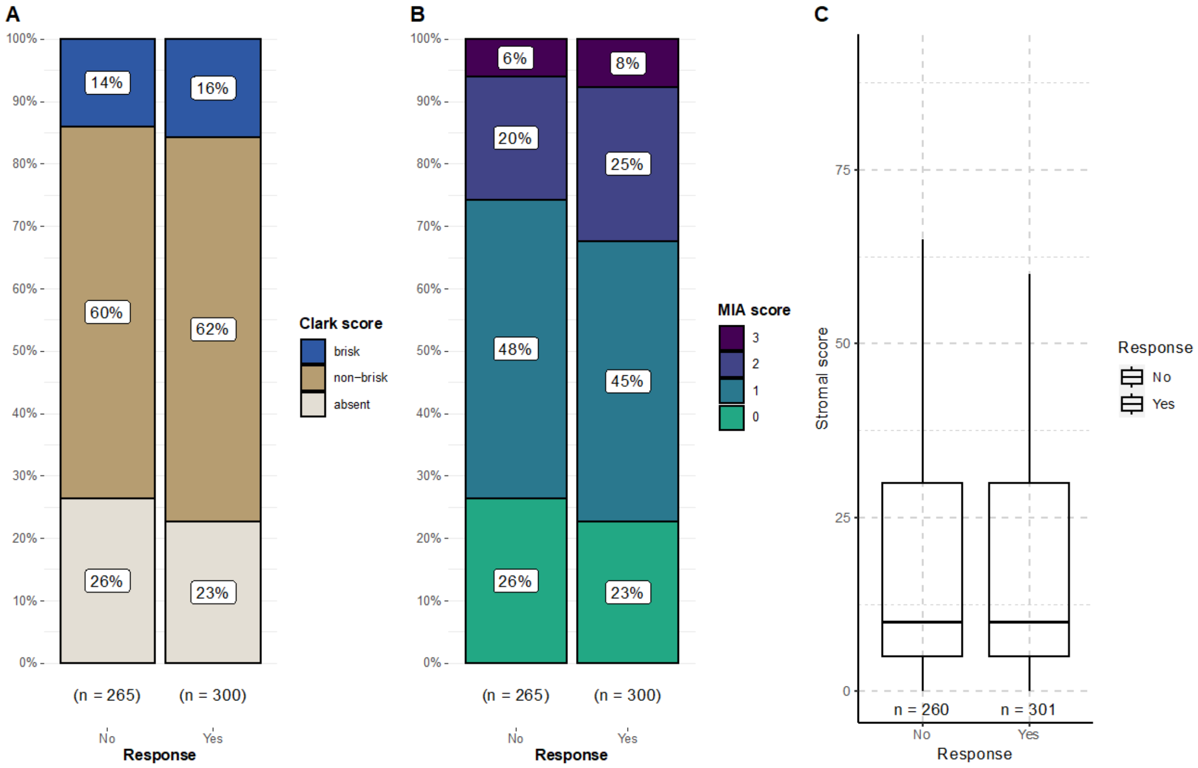

Supplementary figure 2. Kaplan Meier curves for progression-free survival (PFS) and overall survival (OS) for the MIA score and Stromal score in patients with a metastatic melanoma specimen with median survival times and corresponding 95% confidence interval. P-values for median survival times were calculated using logrank tests. In the curves for the MIA score, 19 patients were not included in the PFS analysis, and 15 patients were not included in the OS analysis because of missing survival outcomes. In the curves for the stromal score, 17 patients in the PFS analysis and 14 patients in the OS analysis were not taken into account because of missing survival outcome.

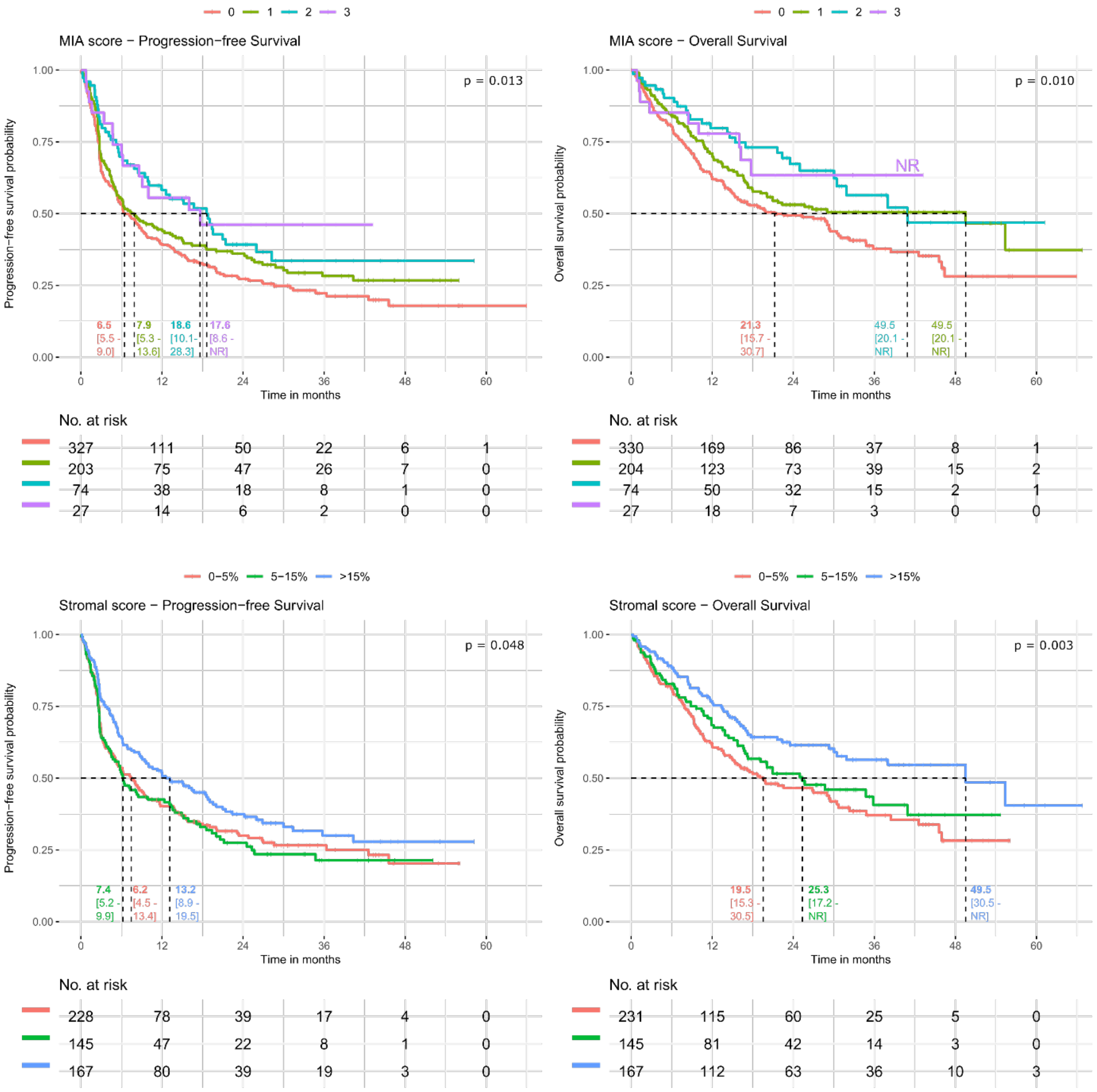

Supplementary figure 3

Heatmap of Clark scores in matched primary and pre-treatment metastatic specimens in 341 patients with advanced cutaneous melanoma treated with ICI. A significant shift towards decreased TIL score from primary to metastatic specimens was found (p < 0.001).

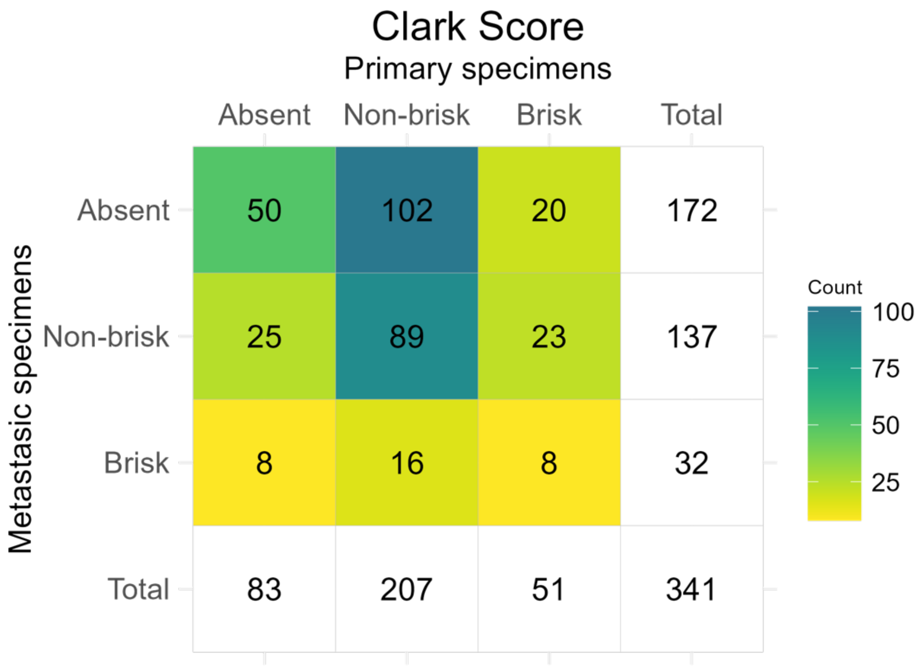


Supplementary figure 4

Heatmap of MIA scores in matched primary and pre-treatment metastatic specimens in 348 patients with advanced cutaneous melanoma treated with ICI. A significant shift in TIL score across the primary and metastatic specimens was found (p < 0.001).

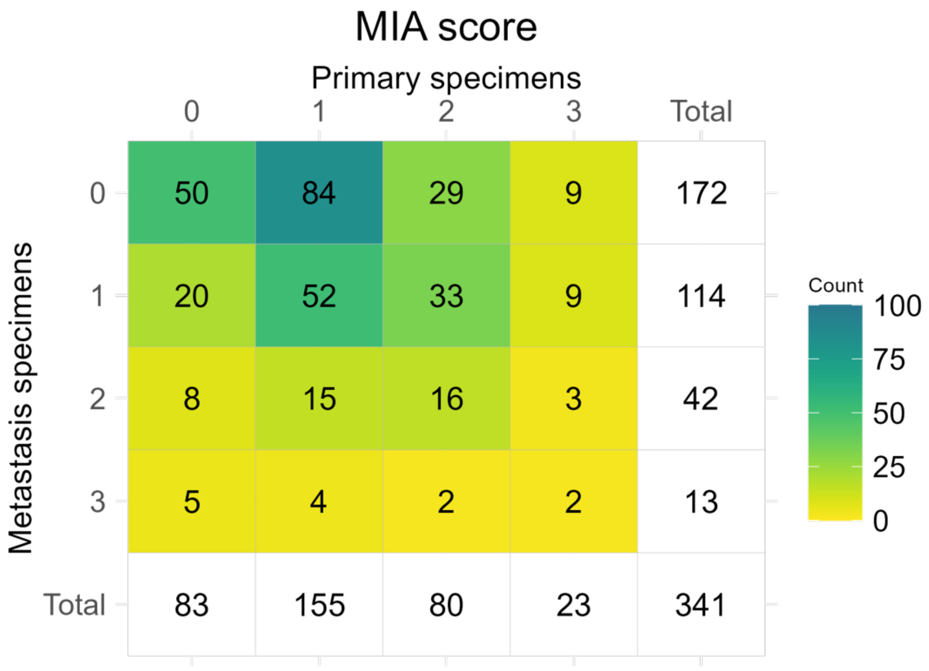

### References


1. Clark, W. H., Jr. *et al.* Model Predicting Survival in Stage I Melanoma Based on Tumor Progression. *JNCI: Journal of the National Cancer Institute* **81**, 1893–1904 (1989).
2. Clemente, C. G. *et al.* Prognostic value of tumor infiltrating lymphocytes in the vertical growth phase of primary cutaneous melanoma. *Cancer* **77**, 1303–1310 (1996).
3. Azimi, F. *et al.* Tumor-Infiltrating Lymphocyte Grade Is an Independent Predictor of Sentinel Lymph Node Status and Survival in Patients With Cutaneous Melanoma. *JCO* **30**, 2678–2683 (2012).
4. Hendry, S. *et al.* Assessing tumor infiltrating lymphocytes in solid tumors: a practical review for pathologists and proposal for a standardized method from the International Immuno-Oncology Biomarkers Working Group. *Adv Anat Pathol* **24**, 311–335 (2017).
